# Performance of temporal artery temperature measurement in ruling out fever: implications for COVID-19 screening

**DOI:** 10.1101/2020.04.24.20070649

**Authors:** Adrian D. Haimovich, R. Andrew Taylor, Harlan M. Krumholz, Arjun K. Venkatesh

**Affiliations:** Yale Department of Emergency Medicine, Yale School of Medicine, 464 Congress St. 260, New Haven, CT, 06516; Yale Department of Internal Medicine, Section of Cardiovascular Medicine, Yale School of Medicine, 333 Cedar Street, New Haven, CT 06520; Yale New Haven Hospital Center for Outcomes Research and Evaluation, 1 Church Street, New Haven, CT 06510

## Abstract

The use of non-invasive temperature testing methods like temporal artery thermometers (TATs) is growing exponentially in the face of the ongoing COVID-19 pandemic. We performed a retrospective analysis of over 1.8 million emergency department electronic health records to identify assess the performance of TAT measurement using patients with near-contemporaneous temperature measurements taken via rectal or oral approaches. Using over 17,000 matched measurements, we show poor fever sensitivity using TAT. We show that sensitivity is significantly improved by lowering the fever threshold and describe limits of agreement between methods of measurement. Our findings suggest that private, public, and healthcare delivery organizations may need to reconsider how we perform high-volume screening during this time of crisis and has implications for return-to-work protocols.

## Background

Current CDC recommendations for mitigation of community COVID-19 transmission include temperature screening.^1^ Port-of-entry symptom and temperature screenings are now common worldwide in community settings such as airports as well as healthcare settings such as Veterans Health Administration Hospitals and are being considered as part of return-to-work protocols.^2, 3^ Due to low cost and ease-of-use, temporal artery thermometers (TATs) applied to the forehead are widely employed to screen for fever, but prior literature has suggested poor sensitivity and high variability.^4, 5^ Published data are limited by small per-study sample sizes and a focus on pediatric, surgical, and intensive care settings that are not generalizable to real-world screening populations.

## Objective

We sought to determine the real-world test performance of TATs for fever rule-out by utilizing a large electronic dataset of emergency department encounters for whom universal temperature screening was conducted as part of standard triage processes. For reference standards, we included rectal temperature, a widely recognized core temperature, as well as oral temperature, which benefits from wide clinical acceptance and robust specificity.^5^ Our primary objective was to determine TAT sensitivities and specificities across a range of temperatures in comparison to rectal and oral cutoffs of 100.4°F/38°C.^4, 5^ Our secondary outcome was limit-of-agreement (LOA) by Bland-Altman analysis.

## Methods and Findings

We extracted temperature measurements and method of temperature assessment from electronic health record (EHR) data (Epic, Verona, WI) collected between March 2013 and June 2019 within a large hospital system comprising ten acute care sites. These data were part of a quality improvement effort and exempted from review by the Yale University Institutional Review Board. We identified paired per-patient data where a TAT measurement was documented within 15 minutes of a rectal temperature measurement or oral temperature. When multiple measurements were taken with a single modality within the defined interval, the mean value was used.

We identified 1.84 million adult (age *>* 18 years) emergency department visits by 602,089 patients with over 4.6 million temperature readings; there were 1,293 paired readings from 1,276 encounters that met our inclusion for TAT versus rectal measurement and 16,132 readings from 16,031 encounters for TAT versus oral measurement (Table 1). Fever prevalence in the rectal and oral temperature populations were 34.4% and 4.3%, respectively. Using a threshold of 100.4°F, TAT measurement identified fever compared with the rectal reference with sensitivity 0.27 (95% CI: 0.27-0.31), specificity 0.98 (0.96-0.99), PPV 0.85 (0.79-0.91), NPV 0.72 (0.69-0.74). TAT measurement identified fever compared with the oral reference with a sensitivity 0.23 (95% CI: 0.20-0.26), specificity 0.99 (0.99-0.99), PPV 0.53 (0.48-0.59), NPV 0.97 (0.96-0.97). We did not observe significant differences in fever sensitivity when limiting our paired analysis to five or ten minute windows. Decreasing the threshold for fever using TAT improved sensitivity with respect to the rectal temperature gold standard of 100.4°F (Figure 1). Using a cutoff of 99°F, for example, increased sensitivity to 0.63 (0.58-0.67) and NPV to 0.82 (0.79-0.84), while specificity decreased to 0.86 (0.83-0.88) and PPV to 0.7 (0.65-0.74). The mean differences and LOA between TAT and rectal temperatures and TAT and oral temperatures were -1.13 (LOA: -6.69 to 4.42)°F, and -0.45 (-3.89-2.99)°F, respectively (Figure 2).

**Table 1:**
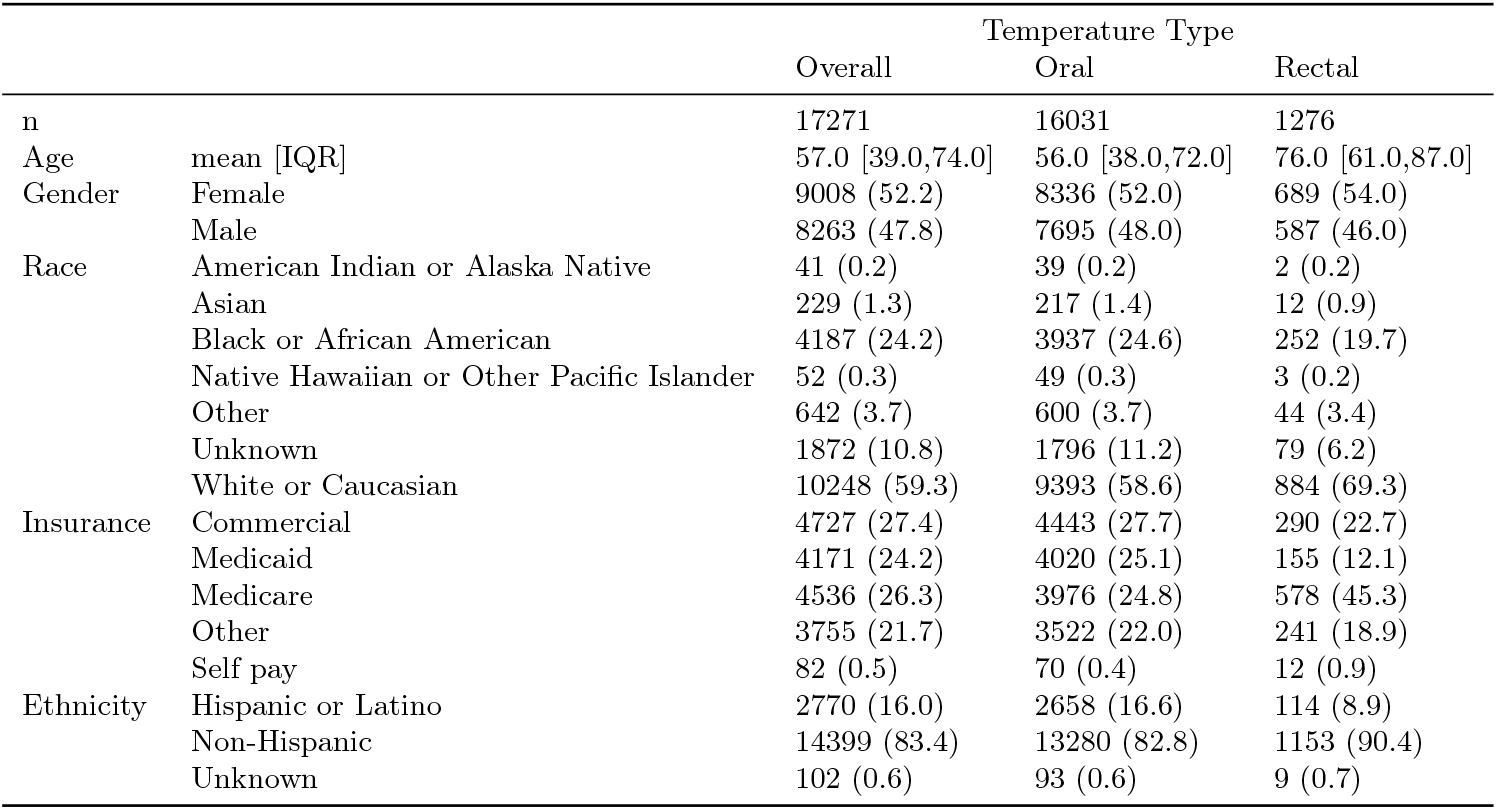
Characteristics of Patients with Temperature Measurement in the ED

**Figure 1:**
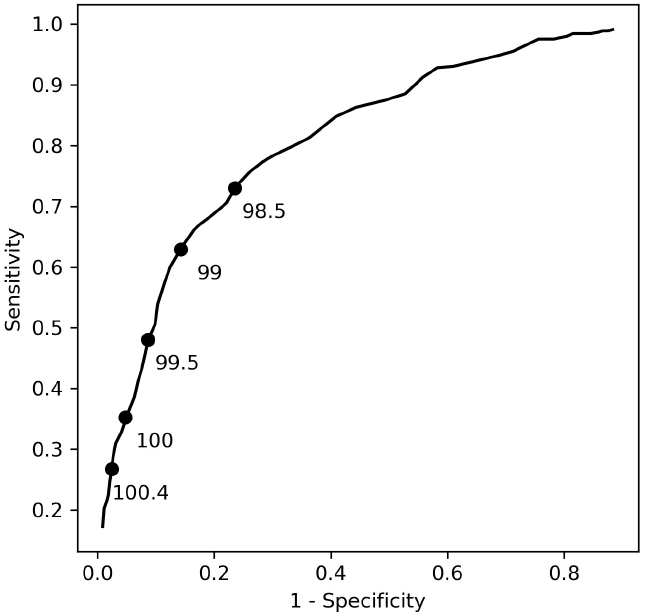
Specificity/sensitivity plot for fever screening by TAT as compared to rectal temperature of 100.4°F. Select temperature cutoffs for TAT are labeled.

**Figure 2:**
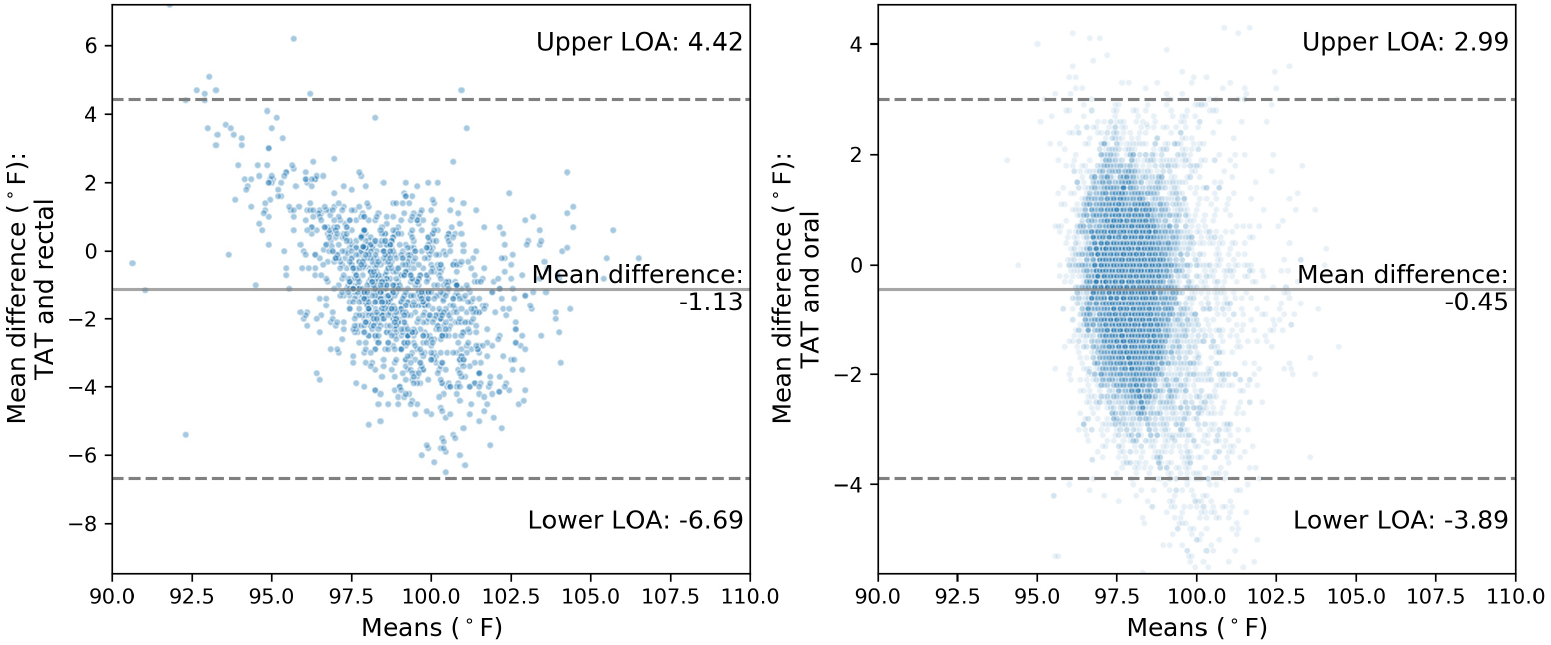
Bland-Altman plots of TAT and (A) rectal and (B) oral measurements. Mean difference is shown with the solid line, limits of agreement as defined by two standard deviations are shown with dashed lines.

## Discussion

To our knowledge, this is the largest study to date comparing TAT measurement to rectal or oral measurements in an adult population. Consistent with prior work,^4, 5^ we found TATs had poor test performance, identifying less than one in three positive cases using either rectal or oral reference measurements. We observed that decreasing TAT temperature thresholds yielded significantly improved test sensitivity with modest losses in specificity. Our analysis is limited by its reliance on selection of patients who had paired measures, which may limit the generalizability. Nevertheless, these data raise questions about the sensitivity of TAT screening for the detection of people with fever amidst the current COVID-19 pandemic. Using a TAT cutoff of 100.4°F, the majority of people who would meet criteria for fever would be wrongly classified as afebrile.

## Data Availability

All authors had full access to the data. The data are available for external use at this time.

